# Variant-specific humoral immune response to SARS-CoV-2 escape mutants arising in clinically severe, prolonged infection

**DOI:** 10.1101/2024.01.06.24300890

**Authors:** Theresa Günther, Michael Schöfbänker, Eva Ulla Lorentzen, Marie-Luise Romberg, Marc Tim Hennies, Rieke Neddermeyer, Marlin Maybrit Müller, Alexander Mellmann, Georg Lenz, Matthias Stelljes, Eike Roman Hrincius, Richard Vollenberg, Stephan Ludwig, Phil-Robin Tepasse, Joachim Ewald Kühn

## Abstract

Neutralising antibodies against the SARS-CoV-2 spike (S) protein are major determinants of protective immunity, though insufficient antibody responses may cause the emergence of escape mutants. We studied the intra-host evolution and the humoral immune response in a B-cell depleted, haemato-oncologic patient experiencing clinically severe, prolonged SARS-CoV-2 infection with a virus of lineage B.1.177.81.

Bamlanivimab treatment early in infection was associated with the emergence of the bamlanivimab escape mutation S:S494P. Ten days before virus elimination, additional mutations within the N-terminal domain (NTD) and the receptor binding domain (RBD) of S were observed, of which the triple mutant S:Delta141-4 E484K S494P became dominant. Routine serology revealed no evidence of an antibody response in the patient. A detailed analysis of the variant-specific immune response by pseudotyped virus neutralisation test (pVNT), surrogate VNT (sVNT), and immunoglobulin-capture EIA showed that the onset of an IgM-dominated antibody response coincided with the appearance of escape mutations. The formation of neutralising antibodies against S:Delta141-4 E484K S494P correlated with virus elimination. One year later, the patient experienced clinically mild re-infection with Omicron BA.1.18, which was treated with sotrovimab and resulted in a massive increase of Omicron-reactive antibodies.

In conclusion, the onset of an IgM-dominated endogenous immune response in an immunocompromised patient coincided with the appearance of additional mutations in the NTD and RBD of S in a bamlanivimab-resistant virus. Although virus elimination was ultimately achieved, this humoral immune response escaped detection by routine diagnosis and created a situation temporarily favouring the emergence of escape variants with known epidemiological relevance.

## Introduction

About four years into the SARS-CoV-2 pandemic, with a death toll of at least seven million people out of about 700 million diagnosed with COVID-19, the WHO declared the end of the health emergency state in May 2023. A rapid succession of viral immune escape variants has led to the dominance of Omicron and its descendants [1]. Hybrid immunity resulting from previous vaccinations and infections appears to contribute to Omicron causing less severe disease than its predecessors [2]. Nevertheless, the high disease burden and lost working hours demand close monitoring of further viral evolution and gaining a deeper insight into the mechanisms of variant formation.

Immunocompromised patients have been put front and centre among several hypotheses explaining the seemingly erratic emergence of escape variants, such as unnoticed spread within distinct human populations and reciprocal transmission between an animal reservoir and humans as reviewed by Markov et al [3]. In the immunocompromised host, chronic infections, inadequate immune responses, and therapeutic measures exerting immunological pressure, like monoclonal antibody monotherapies, insufficiently acting polyclonal COVID-19 convalescent plasma (CCP) or antiviral drugs eliciting mutations, are supposed to alleviate the selection of escape variants [4, 5].

While S-specific neutralising antibodies confer protection against severe disease and, albeit to a lesser extent, against re-infection, they simultaneously contribute to the evolution of escape variants [6-9]. Due to mutations within the antigenic domains of S and recombination events, circulating virus variants efficiently overcome immunity in vaccinees and convalescents and render many therapeutically relevant monoclonal antibodies and CCP ineffective. Importantly, immune imprinting has been discussed to increase selection pressure, thus favouring immune evasion [10]. To better understand the mechanisms of antibody-driven selection processes and, thus, to better predict future developments, it is crucial to investigate the interaction between humoral immune response and infecting virus in immunocompromised patients on a temporal scale and concerning its specificity.

In the case reported here, we analysed the interplay between intra-individual virus evolution and consecutively adapting antibody reaction. Since the patient was closely monitored due to his severely compromised immune system, we were able to study the prolonged SARS-CoV-2 infection and corresponding immune response at high temporal resolution. Our data highlight the importance of the patient’s IgM response in neutralising and eliminating escape variants during the first critical COVID-19 episode. An Omicron infection about one year later was clinically mild and characterised by intense stimulation of IgG and IgA antibodies, mainly targeting cross-reactive epitopes of the first infecting virus strain and the vaccine strain, respectively. Omicron-specific epitopes were targeted to a lesser extent, most likely due to immunological memory and immune imprinting [11, 12]. These results deepen our understanding of escape variant selection in immunocompromised patients and provide methods to monitor endogenous immune responses in prolonged infections in this patient cohort.

## Materials and Methods

### SARS-CoV-2 nucleic acid detection

Viral RNA was extracted from patient samples by the *QIAsymphony DSP Virus/Pathogen Kit* (Qiagen N.V., Hilden, Germany) and reverse transcribed using the *SuperScript^TM^ III Platinum^TM^ One-Step qRT-PCR System* (Invitrogen/Thermo Fisher Scientific, Waltham, MA, USA) on the *QIAsymphony* instrument (Qiagen). In-house quantitative real-time PCR (qRT-PCR) was performed using the *LightMix^®^ Modular SARS-CoV (COVID-19) E-gene* for sarbecovirus detection and the *LightMix^®^ Modular SARS-CoV-2 (COVID19) RdRP-gene* for diagnostic confirmation of SARS-CoV-2 (TIB Molbiol/Roche Diagnostics, Berlin/Mannheim, Germany). FAM label detection assays were performed on a *RotorGeneQ* device (Qiagen).

### Whole genome sequencing

Complementary DNA (cDNA) was transcribed from viral RNA isolated from nasopharyngeal swabs using the *LunaScript^®^ Reverse Transcription Kit* according to the manufacturer’s instructions (New England Biolabs, Ipswich, USA). Whole genome sequencing (WGS) was performed utilising the *EasySeq^TM^ SARS-CoV-2 Whole Genome NGS Sequencing Kit* (NimaGen B.V., Nijmegen, The Netherlands) for multiplex amplicon-based preparation of next generation sequencing (NGS) libraries. Briefly, the amount of cDNA was adjusted to the cycle threshold (ct) value as recommended by the manufacturer (NimaGen B.V.) to perform the amplification and barcoding of samples. Subsequently, samples were prepared for running on the *Illumina MiSeq^TM^* platform using the 150 bp paired-end sequencing chemistry (Illumina Inc., San Diego, CA, USA). The resulting fastQ files were further processed (primer removal, quality trimming) and mapped onto the SARS-CoV-2 reference genome NC_045512.3, and variants (substitutions, smaller and larger insertions or deletions) were extracted using the Ridom *SeqSphere+* software version 9 (Ridom GmbH, Muenster, Germany).

### Routine serology

Routinely, IgG antibodies against the nucleocapsid (N) protein (IgG-N) of SARS-CoV-2 were qualitatively assessed by the commercially available, CE/IVD certified chemiluminescence microparticle immunoassay (CMIA) *Abbott Architect SARS-CoV-2 IgG* according to the manufacturer’s instructions (Abbott Diagnostics, Abbott Park, North Chicago, Illinois, US). Accordingly, IgG antibodies against the SARS-CoV-2 receptor binding domain (RBD) of the S protein subunit S1 (IgG-S) were quantified by the CE/IVD certified CMIA *Abbott Architect SARS-CoV-2 IgG II Quant* (Abbott Diagnostics). Anti-IgG-S values were expressed as arbitrary units (AU)/mL, values greater than or equal to 50.0 AU/mL indicating seropositivity.

To discriminate between N-, S1-and RBD-specific antibodies, the immunostrip assay *recomLine SARS-CoV-2 IgG* (Mikrogen GmbH, Neuried, Germany), which contains recombinant target antigens, was performed. The manufacturer claims a sensitivity of 96.3% and a specificity of 98.8%. For assessment of IgM, immunostrips of the IgG kit were probed with IgM-specific reagents (Mikrogen). Antibody levels were visually determined according to the manufacturer’s guidelines as ordinal values using the cut-off band of immunostrips as an internal reference. Results of individual target-specific bands were rated on an ordinal scale as non-detectable (-), below the cut-off (+/-), with cut-off intensity (+), above the cut-off (++), and very strong intensity (+++).

We applied the CE/IVD certified *cPass^TM^ SARS-CoV-2 Neutralization Antibody Detection Kit* (GenScript Biotech, Mainz, Germany) to assess the neutralisation capacity of patient antibodies. This surrogate virus neutralisation test (sVNT) quantifies the inhibition of wild-type (wt) RBD binding to hACE2 by antibodies in a blocking ELISA format and correlates with infectious virus neutralisation assays [13-15]. Following the manufacturer’s manual, patient samples were diluted 10-fold and measured in technical duplicates. Binding inhibition was calculated as 1 - (OD value of sample/OD value of negative control) × 100 %. Values less than the cut-off of 30 % are considered negative; values at or above the cut-off indicate the presence of SARS-CoV-2 neutralising antibodies.

### Cloning of spike constructs used in pVNT, sVNT and EIA

Substitution S494P was introduced into the vector pCG1-SARS-2-S-Delta1253, which was described by Schoefbaenker et al. [16], by opening the plasmid with BamHI and AgeI and inserting PCR fragments amplified from pCG1-SARS-2-S with primer pairs CG1-S-Bam fwd and CG1-494P bwd, and CG1-494P fwd and CG1-S-Age bwd, respectively. This yielded the vector pCG-SARS-2-S-494P-Delta1253 (**Supplementary Table 1a, b**. Accordingly, vectors pCG1-SARS-2-S-484K-494P-Delta1253 and pCG1-SARS-2-S-Delta141-4-484K-494P-Delta1253 containing the substitution E484K and deletion 141-4 were generated by PCR-mediated site-directed mutagenesis. Primers and constructs are listed in **Supplementary Table 1a, b**. The spike protein with the Omicron BA.1-specific amino acid sequence and a C-terminal 21aa deletion was expressed from vector pcDNA3.1 SARS-2 Omicron comprising a synthetic S-insert (Thermo Fisher Scientific) as described [16].

Cloning of the vectors pEN-secNL-RBD and pEN-secNL-RBD Omicron expressing the secreted form of NanoLuc^®^ luciferase (NLuc, Promega, Walldorf, Germany) fused to the wt RBD and Omicron BA.1 RBD, respectively, were performed according to Schoefbaenker et al. [16] Constructs pEN-secNL-RBD-S494P and pEN-secNL-RBD-484K-494P were generated by opening pEN-secNL-RBD with BamHI and NotI and inserting PCR fragments amplified with primers RBD-BamHI_2 fwd and RBD-NotI bwd from pCG1-SARS-2-S-494P-Delta1253 and pCG1-SARS-2-S-484K-494P-Delta1253, respectively, by InFusion cloning (Takara Bio, Mountain View, CA, USA) (**Supplementary Table 1a, b**). The vector pEN-secNL-RBD-E340K was obtained by opening pEN-secNL-RBD with NheI and NotI and inserting PCR products amplified from pEN-secNL-RBD with primer pairs secNL fwd and wt E340K bwd, and wt E340K fwd and RBD Not1 bwd, respectively, by two-fragment InFusion cloning. The construct pEN-secNL-RBD-Omicron-E340K was generated in the same way using pEN-secNL-RBD Omicron as the target sequence and primers Omicron RBD E340K fwd and bwd to introduce the substitution E340K (**Supplementary Table 1a, b**).

Plasmid pEN-secNL-15-307 expressing the NTD of S fused to secreted NLuc (secNL) was generated as described [16]. Vectors pEN-secNL-NTD-Delta141-4 and pEN-secNL-NTD-Omicron were generated accordingly by inserting PCR products amplified with primers S15-BamH1 fwd and S307-Not1 bwd from pCG-1-SARS-2-S-Delta141-4-484K-494P-Delta1253 and pcDNA3.1 SARS-2 Omicron, respectively.

### Pseudovirus-based virus neutralisation test

SARS-CoV-2 neutralisation was analysed with the SARS-CoV-2 S protein-bearing GFP-expressing vesicular stomatitis virus (VSV) pseudotyped system (VSV-pVNT) as described earlier [16-18]. Vectors used to express wt S and variant forms of S are listed in **Supplementary Table 1b**. Patient sera were tested in the pVNT at 1:20, 1:80, and 1:320 dilutions. Four technical replicates were performed, and the mean was calculated. SARS-CoV-2 S antibody negative and positive sera pools were used as controls. GFP-positive cells were quantified with the Celigo Image Cytometer (Nexcelom/Perkin Elmer Inc., Waltham, MA, USA). The degree of neutralisation was calculated as the reduction of the GFP signal (%) = (1 – GFP-positive cells of the treated sample/GFP-positive cells of the untreated sample) x 100%. The reduction of GFP-positive cells by ≥50% was rated as a positive result in the pVNT [16].

### In-house sVNT and in-house immunoglobulin capture EIA

Expression of NLuc-tagged recombinant proteins by transient transfection of cell cultures and quantification of inhibitory antibodies by the in-house sVNT and immunoglobulin capture EIA were performed as described. Sera were tested in the sVNT at a dilution of 1:20. Luciferase activity in the absence of human serum, and SARS-CoV-2 antibody negative and positive human serum pools, respectively, served as controls. Inhibition of the binding of the RBD to hACE2 was calculated as reduction of the NLuc signal (%) = (1 – NLuc signal of the sample/NLuc signal of the untreated sample) x 100. The cut-off was set at 25% reduction.

To determine IgG, IgA, and IgM antibodies in human sera using secNLuc-tagged spike antigens in a heavy chain-capture EIA, sera were tested at a dilution of 1:100. Values were expressed as Ig-class-specific activity (rlu) subtracted by activity in the absence of serum samples. SARS-CoV-2 positive and negative serum pools served as controls.

## Results

### Patient history

We report a patient in their 60s diagnosed with acute myeloid leukaemia (AML M1, intermediate risk by ELN). They initially received chemotherapy according to the 7+3 regimen plus gemtuzumab ozogamicin, followed by therapy with high-dose cytarabine/gemtuzumab ozogamicin. Due to AML relapse six months later, the first allogeneic stem cell transplant was performed. After ten months, the patient suffered another AML relapse, whereupon a second allogeneic stem cell transplant was performed. As a result, the patient developed graft-versus-host disease in the intestine (grade III), liver (grade II), and skin (grade II). Six months after transplantation, Epstein-Barr virus-induced lymphoproliferation was detected, necessitating B-cell depleting therapy with rituximab.

### The clinical course of SARS-CoV-2 infection

When the patient was admitted to the hospital two months later for *Legionella* pneumonia and consecutive cardiac decompensation, the AML was in remission. During hospitalisation, the patient developed coughs and progressive dyspnoea. SARS-CoV-2 infection was confirmed by qRT-PCR in a nasopharyngeal swab (defined as day 1 of the disease) (**Fig. 1a**). On admission to the isolation ward, laboratory analysis revealed leukopenia (2.420/µL) with 90.4 % neutrophils and marked lymphopenia (1.7 % lymphocytes). Immunoglobulins were markedly decreased (IgG 458 mg/dL), and CD19+ B lymphocytes were almost completely depleted at 0.2 % (1 cell/µL absolute). CD4+ T lymphocytes (25.6 %, 81 cells/µL) were significantly reduced, while CD8+ T lymphocytes were within the normal range at 85.3 %.

**Fig. 1:**
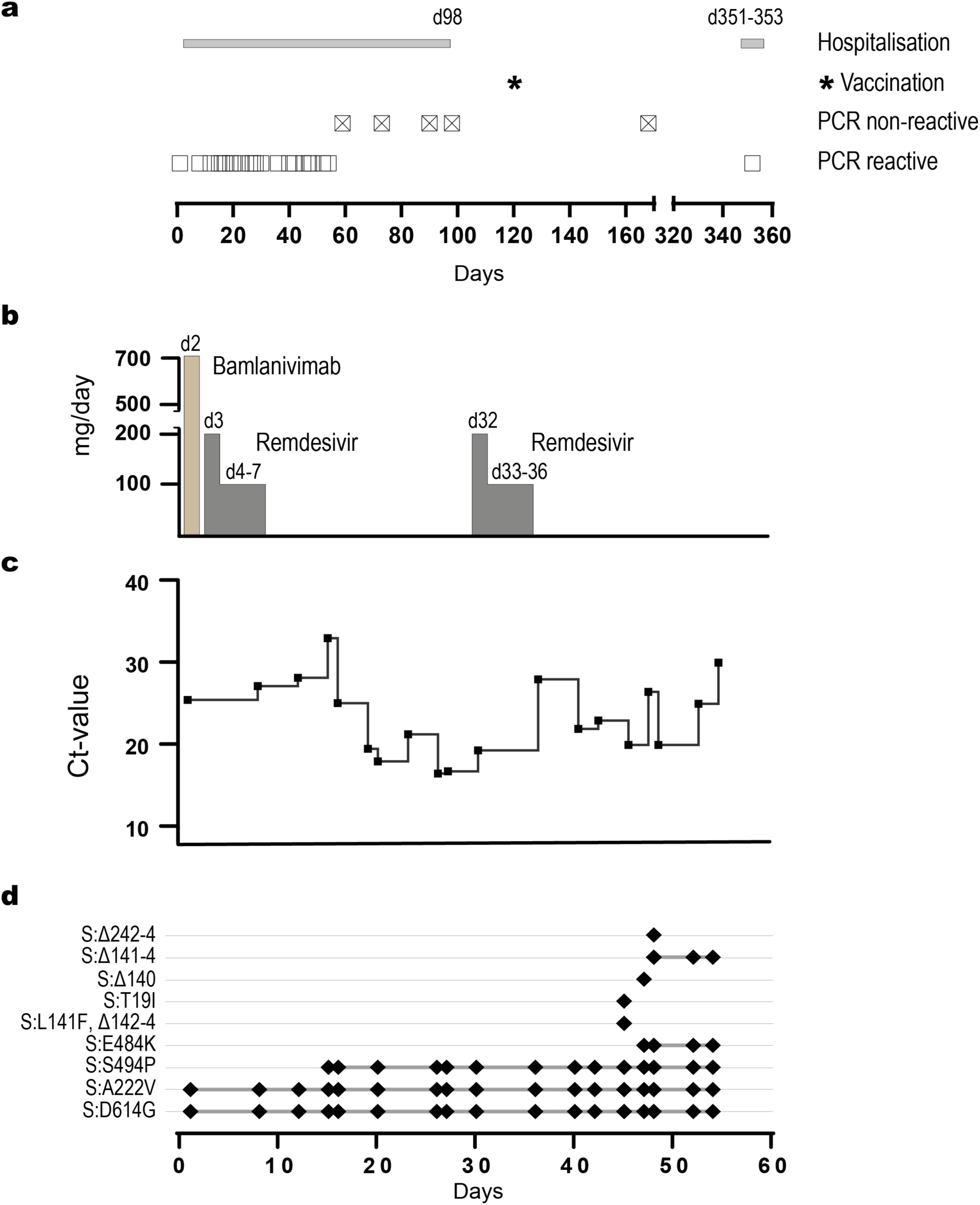
Course of the first and second episode of SARS-CoV-2 infection. **(a)** Duration of hospitalisation (grey bar), time point of first vaccination (asterisc) and detection of SARS-CoV-2 RNA in nasopharyngeal swabs by qRT-PCR. Positive and negative PCR results are indicated by squares and crossed-out squares, respectively. The day of sampling after the initial diagnosis (day 1) is indicated. **(b)** Time course of the administration and dosage of antiviral therapeutics; the timeline of sampling is indicated in panel d. **(c)** Relative viral load in nasopharyngeal swabs during the first episode as measured by Ct values of qRT-PCR, for the timeline of sampling see panel d. **(d)** Mutations detected in S during the first episode. The repeated detection of mutations in consecutive samples is marked with a grey line. GISAID accession numbers of viral genome sequences are given in Supplementary Table 2. The day of sampling after the initial diagnosis (day 1) is indicated.

Following the COVID-19 treatment guidelines at the time, the patient received monotherapy with the S-specific monoclonal antibody bamlanivimab (700 mg) on day 2. In addition, they were treated with remdesivir for five days (200 mg on day 3 and 100 mg on days 4 to 7) (**Fig. 1b**). Upon initiation of treatment, dyspnoea declined, whereas fatigue and intermittent diarrhoea persisted. On day 19 fever and coughs reoccurred, while the persisting fatigue worsened. Again, remdesivir was administered for five days (days 32 to 36). The patient tested negative for SARS-CoV-2 on days 56 and 57. Intermittent diarrhoea persisted. The first vaccination against SARS-CoV-2 was carried out with Spikevax^®^ (mRNA 1273, Moderna) on day 122. On day 351, the patient experienced a second episode of COVID-19 and received sotrovimab (500mg) on day 352 (**Fig. 1a**). Re-infection resulted in a rapidly self-limiting disease associated with mild symptoms, e.g., headache, myalgia, rhinorrhoea and loss of taste.

The patient was monitored in short intervals for the presence of SARS-CoV-2 RNA in nasopharyngeal swabs by qRT-PCR. On day 1, a cycle threshold (Ct) value of approx. 25 was determined, which corresponds to a viral load of roughly 10^6^ copies of SARS-CoV-2 RNA/mL. Upon commencement of bamlanivimab and remdesivir treatment, RNA levels decreased during the first two weeks. Viral load in swabs reached levels close to the detection limit of PCR on day 15 (**Fig. 1c**). Starting on day 16, viral load in swabs strongly increased and peaked around days 26 and 27. The second remdesivir therapy was associated with a transient reduction of viral RNA levels between days 30 and 36. After completion of remdesivir treatment, the second rise in viral RNA levels was observed, peaking on days 45 and 48. At later time points, RNA levels of nasopharyngeal swabs strongly decreased, and the last positive sample was acquired on day 54 (**Fig. 1c**). In swabs collected between days 73 and 168 viral RNA was not detectable. On day 351, SARS-CoV-2 re-infection was diagnosed by two independent point-of-care tests, i.e., ID NOW^TM^ COVID-19 (Abbott) and BioFire^®^ Respiratory Panel 2.1 *plus* (bioMérieux, Nürtingen, Germany). High levels of SARS-CoV-2 RNA (Ct value 13.14) were found in a nasopharyngeal swab on day 352 (**Fig. 1a**).

### Sequencing results

The significant increase in RNA levels, starting 16 days after disease onset, indicated virological failure of the combined bamlanivimab and remdesivir therapy. This prompted us to search for escape mutations by whole genome sequencing of viral RNA from nasopharyngeal swabs. Sequence analysis on day 1 revealed that the patient had initially been infected with SARS-CoV-2 of PANGOLIN lineage B.1.177.81 (20E, EU1) [19], a variant that became rapidly dominant in Europe during the summer of 2020 [20]. Within the S gene typical amino acid (aa) exchanges of this lineage were found (S:A222V, S:D614G) (**Fig. 1d, Supplementary Table 2**). On day 15, i.e., shortly before the first relapse, the exchange S:S494P within the RBD of the S protein was revealed (**Fig. 1d**). Additional substitutions in the S protein were not observed. Variant virus carrying solely the S:S494P substitution dominated from days 15 to 42.

Coinciding with the second, apparently, biphasic peak in viral RNA levels, variant S:S494P acquired several additional mutations within the NTD and the RBD of S, respectively. On day 45, deletion S:Delta141-3 and substitution S:Y144F were confirmed in two independent samples. Substitution S:T19I was detected in one sample from day 45 but not found at later time points (**Fig. 1d, Supplementary Table 2**). Sequencing on day 47 indicated deletion S:Delta140. Deletion S:Delta141-144 was detected in all sequences from day 48 to 54. One out of two individual sequencing reactions from day 48 indicated the deletion S:Delta242-244. In addition, substitution C1473S in the non-structural protein 3 (NSP3) was detected in this sequencing reaction (EPI_ISL_1643829, **Supplementary Table 2**). Within the RBD, substitution S:E484K was first discovered on day 47 and persisted until virus elimination (**Fig. 1d, Supplementary Table 2**). Sequencing of the SARS-CoV-2 RNA from the swab obtained on day 352 confirmed re-infection with the SARS-CoV-2 variant Omicron BA.1.18 (**Supplementary Table 2**).

### Humoral immune response

Close monitoring of antibody reactivity in peripheral blood samples enabled a detailed analysis of the humoral immune response to both infection episodes. A total of 17 serum samples were available for serological testing. Eight samples were obtained during the first episode of infection (days 2, 8, 17, 32, 40, 45, 49, and 56). Two sera were collected after virus elimination (days 67 and 90), and two sera were taken again after the first vaccination (days 131 and 138). Finally, five sera were obtained before (day 326), during (days 352 and 353) and after re-infection with Omicron BA.1.18 (days 381 and 391), respectively.

### Routine serology

#### Chemiluminescence microparticle immunoassay (CMIA)

In the quantitative, RBD-specific SARS-CoV-2 CMIA (Abbott), IgG antibodies were absent in the first blood sample obtained on day 2, reached their highest levels on day 8 after administration of bamlanivimab, and slowly decreased afterwards. The first vaccination on day 122 did not significantly increase antibody levels on days 131 and 138, respectively (**Fig. 2a**). When re-infection with the Omicron BA.1 variant was diagnosed on day 352, antibody levels were slightly lower as compared to day 138. Administration of 500 mg sotrovimab on day 353 resulted in a strong increase in RBD-specific IgG antibodies. Comparably high levels of RBD-specific IgG levels were observed on days 381 and 391. IgG antibodies against the nucleocapsid (N) protein were not detected in CMIA (Abbott) until day 381 (data not shown).

**Fig. 2:**
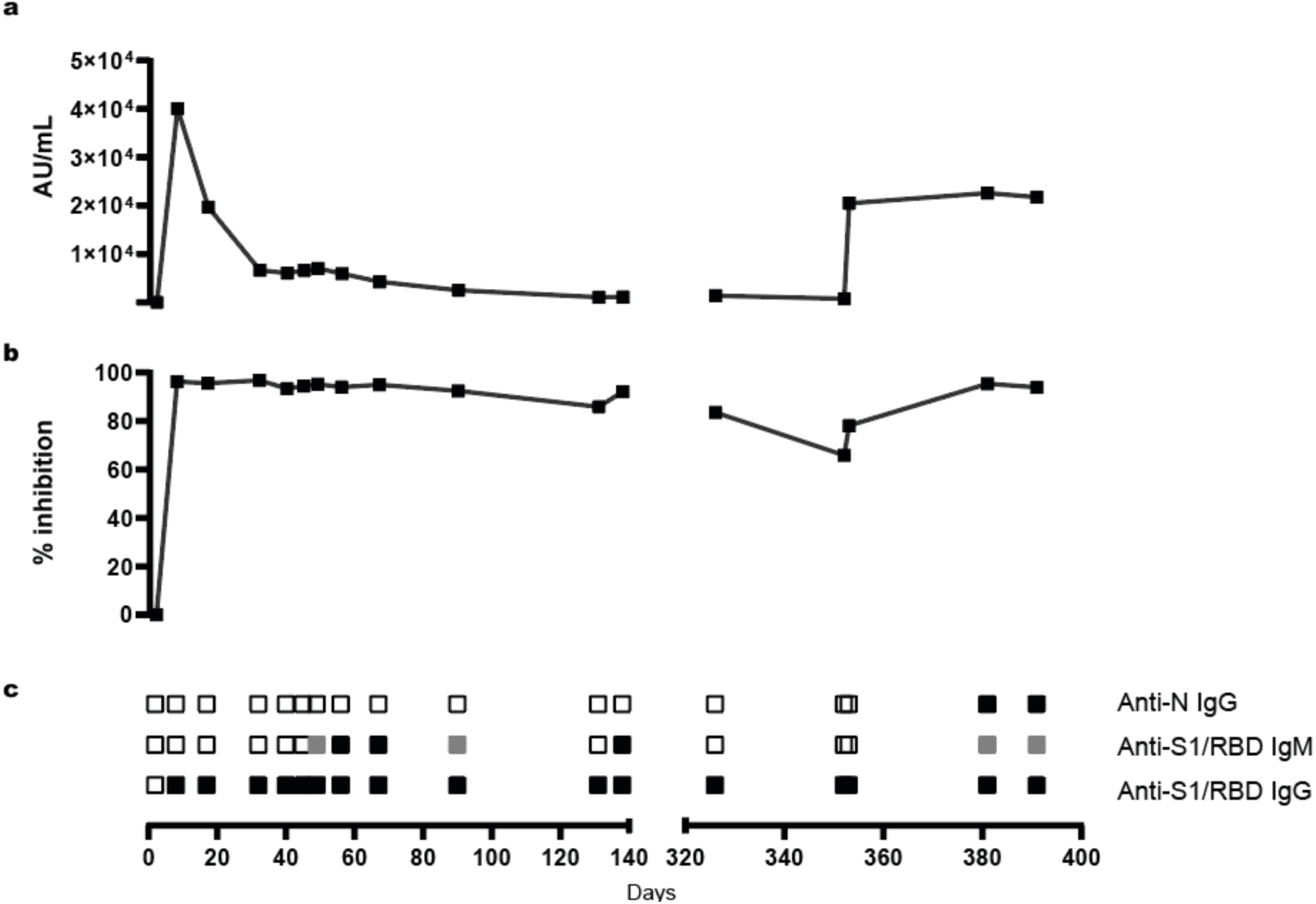
Outcome of routine serological tests. Peripheral blood samples obtained from the patient on the days indicated below panel **c** were tested by the quantitative Abbott Anti-RBD-CMIA **(a)** and the GenScript cPass sVNT **(b)**. Antibody levels detected by Anti-RBD-CMIA and sVNT are given as arbitrary units (AU)/mL, and signal reduction (% inhibition) as compared to the negative control. The sVNT was performed with technical duplicates. **(c)** Qualitative results of the Mikrogen line blot testing for Anti-N IgG, as well as Anti-S1 and Anti-RBD IgM and IgG, respectively, are indicated as follows: open squares: non-reactive, grey squares: reactive below cut-off, black squares: reactive above cut-off.

#### sVNT

The commercial surrogate virus neutralisation test cPass^TM^ (GenScript Biotech) was used to quantify levels of antibodies inhibiting the binding of the RBD to ACE2 [15, 21]. As seen in the quantitative S EIA and line blot, the patient tested negative on day 2. Administration of bamlanivimab resulted in high levels of inhibitory antibodies, which slowly decreased until day 131. The effect of vaccination was reflected by a small increase in inhibitory antibodies on day 138 (**Fig. 2b**). Administration of sotrovimab on day 352 resulted in a moderate increase in antibody levels in sVNT, whereas high levels of inhibitory antibodies were detected on days 381 and 391.

#### Line blot

Qualitative analysis of the IgG response by the Mikrogen line blot assay confirmed the results of the CMIA. SARS-CoV-2-specific antibodies were absent before the administration of bamlanivimab, whereas strong IgG reactivity with S1 and RBD was observed in all samples obtained at later times. N-specific IgG antibodies were only detected on days 381 and 391. Determination of SARS-2-specific IgM antibodies by line blot indicated the presence of IgM antibodies against S1 and RBD on days 49 to 90, day 138 and days 381 and 391 (**Fig. 2c**).

Overall, the results of routine serology suggested the onset of an endogenous humoral immune response at the end of the first infection period and immune responses to vaccination and re-infection with the Omicron variant. This prompted us to better characterise the patientߣs humoral immune response in various S-specific in-house assays with an emphasis on neutralising and/or variant-specific antibodies.

### VSV pVNT

Levels of neutralising antibodies against wt S and S variants, respectively, were quantified by VSV pVNT in blood samples serially diluted 1:20, 1:80, and 1:320. Vectors used to express wt S and variant forms of S are listed in **Supplementary Table 1b**. Using wt S, the reduction of GFP-positive cells by ≥50% was rated as a positive result in the pVNT. During the first episode of infection, S-specific neutralising antibodies were not detected before administration of bamlanivimab, which resulted in high neutralising antibody titres (≥ 1:320) against wt S. Neutralising antibodies reactive with variants S:S494P, S:E484K S494P, and S:Delta141-4 E484K S494P started to rise on days 40, 45 and 49, respectively, and reached 50% inhibition at a serum dilution of 1:20 on days 49 and 56, respectively. As compared to wt S-specific antibodies, levels of these antibodies were significantly lower, peaked on day 56, and then decreased again. On day 56, titres of neutralising antibodies against S:S494P were approximately 4-fold and 16-fold higher as compared to S:E484K S494P and S:Delta141 4-E484K S494P, respectively. Neutralising antibodies against S Omicron (BA.1) were not detected during the first episode of SARS-CoV-2 infection (**Fig. 3a-e**).

**Fig. 3:**
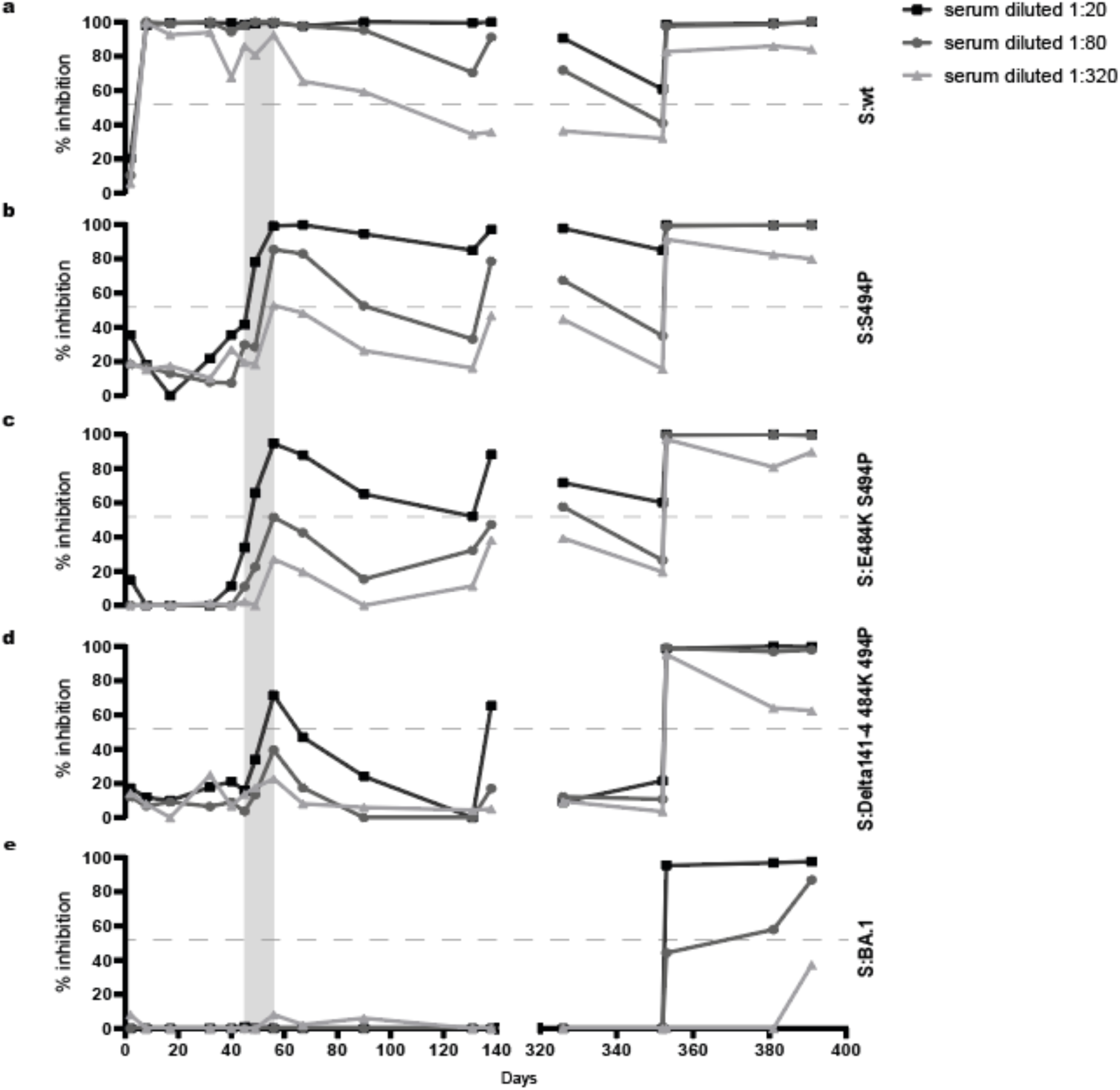
Detection of neutralising antibodies by in-house pVNT. Reactivity of the patient’s sera with wt S **(a)**, S:S494P **(b)**, S:E484K S494P **(c)**, S:Delta141-4 E484K S494P **(d)** and S Omicron BA.1 **(e)** was determined by in-house pVNT. Results are means of four technical replicates and are given as reduction of the GFP signal (%) as compared to the untreated control. Sera were tested at a dilution of 1:20 (black squares), 1:80 (grey squares), and 1:320 (grey triangles). The cut-off of pVNT for wt S was set to ≥50% inhibition (dotted line). Values < 0% were set to zero. The period between the first detection of additional mutations in the NTD and RBD in variant S:S494P on day 45 and virus elimination on day 56 is shaded in grey. The day of sampling after the initial diagnosis (day 1) is indicated below panel **e**.

Administration of the first vaccine dose on day 122 resulted in increased antibody levels against wt S, S:S494P, S:E484K S494P, and S:Delta141-4 E484K S494P on day 138. Neutralising antibodies reactive with S Omicron BA.1 were not elicited. Approximately 4 weeks before (day 326) and immediately upon the onset of the second infection episode (day 352), neutralising antibodies against wt S and variants S:S494P and S:E484K S494P were detected, whereas no reactivity was found with S:Delta141-4 E484K S494P and S Omicron BA.1, respectively. Administration of sotrovimab on day 353 led to strong reactivity with wt S and all variants tested including S Omicron, albeit the increase in S Omicron BA.1-reactive neutralising antibody titres was significantly lower. During the follow-up period neutralising antibodies against S Omicron BA.1 increased from day 381 to day 391. Levels of antibodies against wt S, S:S494P, S:E484K S494P, and S:Delta141-4 E484K S494P remained largely unchanged or decreased.

### In-house sVNT

RBD-fragments N-terminally tagged with secNLuc were used as antigens in the in-house sVNT to detect variant-specific antibodies inhibiting binding to ACE2. The course of inhibitory antibodies directed against wt RBD closely followed the results obtained in the cPass sVNT (**Fig. 2b**). This showed that the administration of bamlanivimab resulted in high levels of inhibitory antibodies, which slowly declined during the first episode of infection. Vaccination moderately increased antibody titres against wt RBD. Wt RBD-specific inhibitory antibodies were also detected on day 326 before Omicron re-infection, increased on day 353 due to the administration of sotrovimab, and remained at high levels on days 381 and 391 (**Fig. 4a**).

**Fig. 4:**
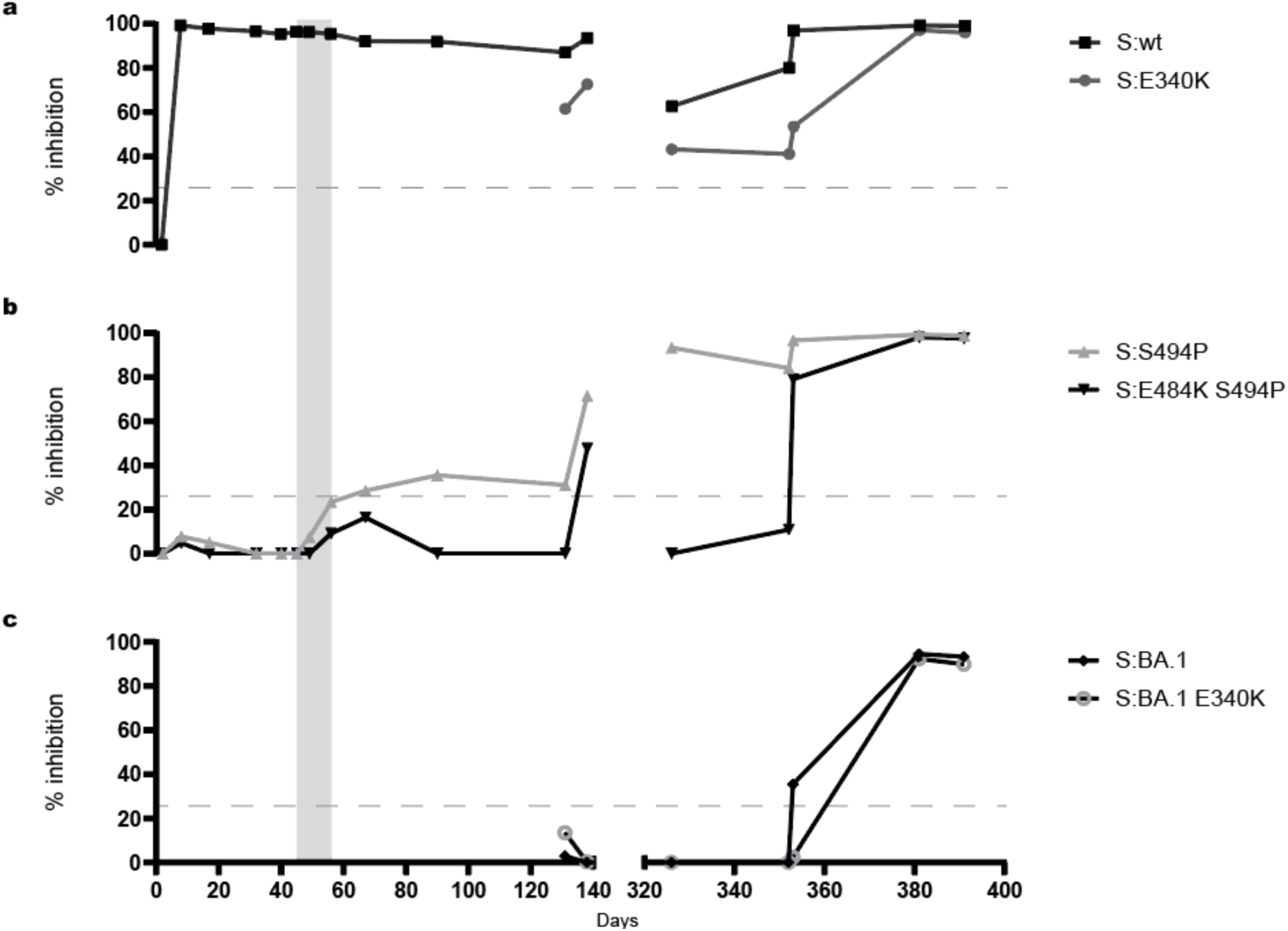
Detection of inhibitory antibodies by in-house sVNT. Reactivity of patient sera with wt RBD and RBD E340K **(a)**, RBD S494P and RBD E484K S494P **(b)**, and RBD Omicron BA.1 and RBD Omicron BA.1 E340K **(c)** was determined by sVNT using N-terminally secNLuc-tagged RBD antigens. Sera were tested at a dilution of 1:20. The cut-off value of the sVNT for wt RBD was set to ≥25% inhibition (dotted line). Values < 0% were set to zero. The period between the first detection of additional mutations in the NTD and RBD in variant S:S494P on day 45 and virus elimination on day 56 is shaded in grey. The day of sampling after the initial diagnosis (day 1) is indicated below panel **c**.

The binding of RBD S494P and RBD E484K S494P, respectively, to ACE2 was not significantly inhibited by the administration of bamlanivimab, highlighting the specificity of the in-house sVNT. An increase in inhibitory antibodies against RBD S494P was observed from days 49 to 56. Antibody levels above the cut-off of the in-house sVNT defined for wt RBD in a vaccination study, i.e., 25 % inhibition as described by Schoefbaenker et al. [16], were first detected on day 67 and remained above the cut-off level in all serum samples taken at later time points. Inhibitory antibodies against RBD E484K S494P did not reach the cut-off of 25% inhibition during the first episode but showed an increase between days 56 and 67 (**Fig. 4b**).

Vaccination increased inhibitory antibody levels against RBD S494P and to a lesser extent against RBD E484K S494P. On day 326, before re-infection with Omicron, inhibitory antibodies against RBD E484K S494P were not detected. Administration of sotrovimab resulted in a strong increase in sVNT titres against RBD S494P and RBD E484K S494 (**Fig. 4b**). The presence of inhibitory antibodies reactive with RBD Omicron BA.1 was determined in response to vaccination and re-infection. RBD Omicron BA.1-reactive inhibitory antibodies were not detected in sVNT until the administration of sotrovimab, which only moderately increased antibody levels. On days 381 and 391, high levels of RBD Omicron BA.1-reactive antibodies were found, indicating the boosting of inhibitory antibodies by the Omicron re-infection (**Fig. 4c**).

To reduce the effect of sotrovimab on the reactivity of serum samples in the sVNT, the substitution S:E340K associated with high-level resistance against sotrovimab was introduced into wt RBD and RBD Omicron BA.1 [22, 23]. In contrast to RBD constructs lacking E340K, administration of sotrovimab on day 353 had only a minor or no effect on antibody reactivity with RBD E340K and RBD Omicron E340K in sVNT, respectively (**Fig. 4a, c**). Subsequently, a strong increase of inhibitory antibodies against RBD 340K and RBD Omicron BA.1 between days 353 and 381 was observed, which further corroborates the triggering of an inhibitory antibody response by Omicron re-infection.

### Immunoglobulin class capture EIA

To delineate the pattern of RBD-specific IgM, IgA, and IgG antibody responses, respectively, secNLuc-tagged RBD-fragments were additionally applied as diagnostic antigens in an in-house Ig class capture EIA. During the first episode of infection, a prominent IgM response against wt RBD, RBD S494P, and RBD E484K S494P peaking around day 56 to 67 was detected. The strongest IgM signal in EIA was observed with wt RBD and RBD S494P, reactivity with RBD E484K S494P was lower. In contrast, RBD-specific IgA responses or an IgG response against RBD S494P and RBD E484K S494P were not observed. As expected, administration of bamlanivimab resulted in very high IgG reactivity with wt RBD on day 8 followed by exponentially decreasing antibody levels between day 8 and 90 (**Fig. 5a-c**). The first vaccination boosted IgM and IgA antibodies reactive with wt RBD, RBD S494P, and RBD E484K S494P, however, did not result in a significant increase in RBD-specific IgG antibodies. IgM, IgA, and IgG antibodies reactive with RBD Omicron BA.1 were not induced by vaccination (**Fig. 5a-c**).

**Fig. 5:**
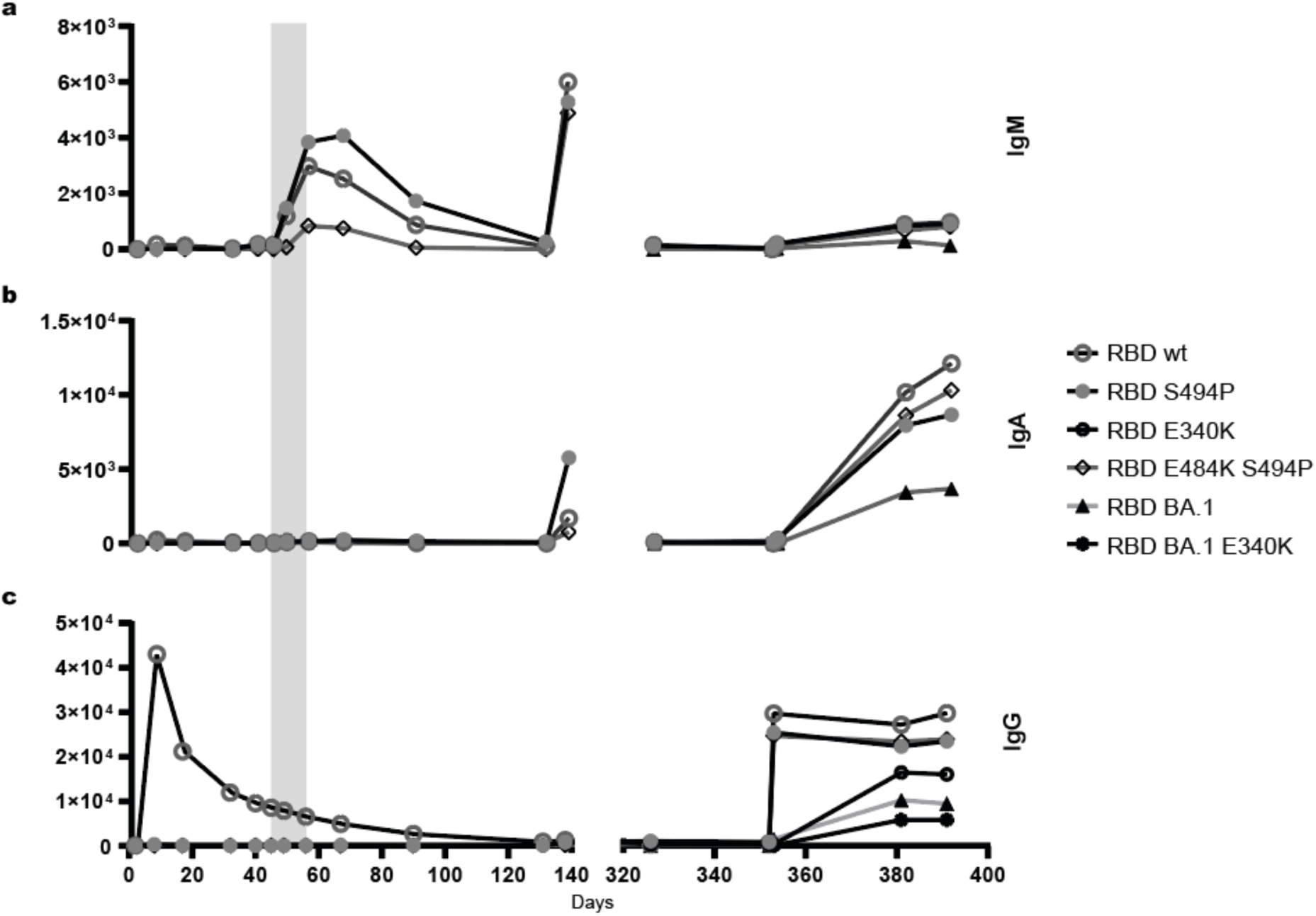
Serum reactivity in the Ig class capture RBD-EIA. IgM reactivity **(a)**, IgA reactivity **(b)**, and IgG reactivity **(c)** of the patient’s sera. Antibody reactivity against wt RBD, RBD S494P, and RBD E484K S494P was determined from days 2 to 391, and reactivity against RBD Omicron BA.1 from days 131 to 391, respectively. Additionally, IgG reactivity with RBD E340K and RBD Omicron BA.1 E340K was tested from days 131 to 391 **(c)**. Sera were analysed at a dilution of 1:100. Signal strength is given in relative light units (rlu). Values < 0 rlu were set to zero. The period between the first detection of additional mutations in the NTD and RBD in variant S:S494P on day 45 and virus elimination on day 56 is shaded in grey. The day of sampling after the initial diagnosis is indicated below panel **c**.

In contrast to the first infection, re-infection with the Omicron variant resulted in low IgM and strong IgA responses, which were dominated by antibodies reactive with wt RBD, RBD S494P, and RBD E484K S494P, however, also contained lower levels of antibodies reactive with RBD Omicron. The prominent IgG reactivity with wt RBD, RBD S494P, and RBD E484K S494P on days 353 to 391 appeared to be mainly caused by administration of sotrovimab. The effect of sotrovimab was much less pronounced on IgG reactivity with RBD Omicron BA.1. A rise in IgG reactivity with RBD Omicron BA.1 from day 353 to 381 indicated the onset of an endogenous RBD-specific IgG response. This was confirmed using the sotrovimab escape-mutants RBD E340K and RBD Omicron E340K as diagnostic antigens. With both antigens IgG levels increasing from day 353 to 381 were observed (**Fig. 5a-c**).

Finally, antibody reactivity with secNLuc-tagged NTD fragments of wt S and variants S:E484K-S494P-Delta141-4 and S Omicron BA.1 was tested in the Ig class capture EIA (**Fig. 6a, b**). Neither during the first episode of infection nor after the first vaccination were significant levels of NTD-specific IgG, IgM, and IgA antibodies detected. In contrast, re-infection with Omicron induced reactivity of NTD-specific IgG and IgA antibodies on days 381 and 391. Levels of IgG antibodies against wt NTD and NTD-Omicron were higher as compared to antibodies against NTD-Delta141-4. Significant IgM responses against the NTD were still not observed (data not shown).

**Figure 6:**
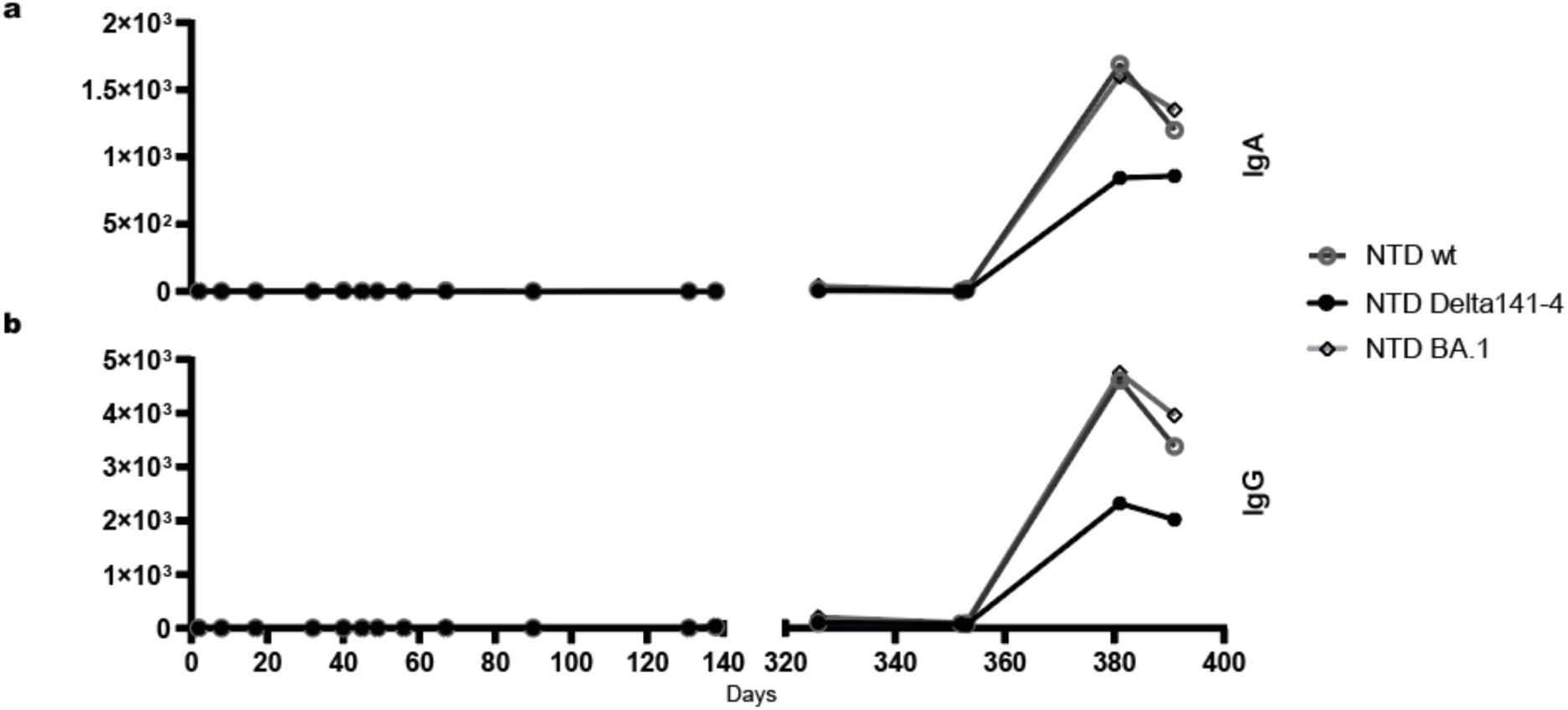
Serum reactivity in in-house Ig class capture NTD-EIAs. **(a)** IgA reactivity, **(b)** IgG reactivity. Sera were tested at a dilution of 1:100. Antibody reactivity is given in relative light units (rlu). Values < 0 rlu were set to zero. The day of sampling after the initial diagnosis is indicated below panel **b**.

## Discussion

Neutralising antibodies against the SARS-CoV-2 spike (S) protein are important determinants of protective immunity [6-8], though passively transferred immunoglobulins or insufficient endogenous antibody responses have been shown to act as driving forces of viral intra-host evolution during chronic infection [24-27]. This Janus-faced role of antibodies in the immunocompromised host became obvious in the case reported here.

After initially satisfactory therapeutic response of a virus of the sublineage B.1.177.81 (clade 20E EU.1) [20] to combined therapy with remdesivir and bamlanivimab, the escape variant S:S494P emerged and again caused a strong increase in viral levels [28, 29]. Other substitutions associated with bamlanivimab escape such as L452, E484, G485, F490 and Q493 have not been observed during the first six weeks of infection [30-35]. In the absence of substitutions increasing the person-to-person spread of SARS-CoV-2, the acquisition of resistance against specific monoclonal antibodies due to single, defined substitutions may be less problematic from an epidemiological point of view [36].

In contrast, insufficient polyclonal antibody reactivities have been described to induce the emergence of variants carrying multiple escape mutations [22, 27, 37], resulting in transmission [38]. Accordingly, the onset of the IgM antibody response in our patient six to seven weeks after infection was associated with the emergence of virus variants harbouring various additional mutations within the NTD and RBD of S. Of note, variants carrying substitutions and deletions at positions 140-4 and 241-3 of the NTD and substitution E484K within the RBD appeared, of which the triple mutant S:Delta141-4 E484K S494P became stably established until virus elimination. The appearance of these variants coincided with temporary and short-lived increases in viral load.

Substitutions at position 484 within the RBD such as E484K occurred in different variants of concern (VOCs) and variants of interest (VOIs), are located in an epitope recognised by most individuals with anamnestic infection and vaccination, cause partial resistance to convalescent sera and confer resistance to bamlanivimab and other class 2 neutralising monoclonal antibodies [22, 30, 31, 33, 35]. The combination of S:E484K with S:S494P has been described as synergistically increasing escape from neutralisation by convalescent sera [29]. In the Omicron subvariants BA.2.10.4, BN.1.1.1, BQ.1.1.11, BQ.1.1.12, BQ.1.19, BY.1.2.2, and CA.2, S:S494P confers growth advantages [39].

Escape mutations within the NTD interfere with the binding of neutralising antibodies to the so-called supersite [25, 31, 40-42], and frequently result from short in-frame deletions clustering in the RDRs (repeated deletion regions) [43]. Deletions similar to S:Delta141-4 observed in the case reported here are found in VOCs B.1.1.7 (alpha) (S:Delta144), B.1.351 (beta) (S:Delta242-4), and B.1.1.529 (Omicron) (S:Delta143-5) and affect loop N3 (residues 141 to 156) as part of the NTD supersite [40]. Dynamic differences in the spectrum of mutations detected in the late phase of infection most likely indicate the presence of a polymorphic virus population occupying discrete niches [36]. This compartmentalisation may have contributed to the poor therapeutic effect of the second remdesivir treatment [44]. In addition, in individuals with prolonged infection, nucleoside analogues like remdesivir may serve as potential co-factors of the emergence of escape mutants by increasing the pool of selectable S variants [45].

In general, there is a trade-off between specific escape properties and indispensable functions of S protein mutants such as binding to the ACE2 receptor with high affinity and/or increased person-to-person spread [36]. Although these aspects might be less relevant in the variants we observed here, there seems to be a trend towards deletions in the NTD and re-occurrence of substitution S:S494P in actual variants [39]. This may point towards the direction of potential escape mutations in future variants. A better understanding of the virus-host interplay resulting in the emergence of escape mutants requires close monitoring of the selective pressure exerted by antibodies during infection.

Administration of therapeutic monoclonal antibodies or CCP, pre-existing humoral immunity, and/or rapidly changing seroreactivity over time may massively impede the assessment of antibody reactivity in the immunocompromised host. Accordingly, in the case reported here, routine serology largely failed to detect unequivocally patient-specific antibody responses against the background of seroreactivity caused by therapeutical monoclonal antibodies. A partial exception was the SARS-CoV-2 line blot [46], which was run in an in-house IgM-specific modification.

We thus established and employed a broad spectrum of variant-specific in-house serologic assays to characterise the humoral immune response in-depth. By this approach onset of the patient-specific neutralising antibody response against S:S494P in pVNT around days 40 to 45 was identified to trigger the emergence of various additional escape variants evolving from the previously dominant bamlanivimab-resistant variant harbouring solely the S:S494P mutation. The increase in S:Delta141-4 E484K S494P-neutralising antibodies in pVNT up to 50% inhibition at a serum dilution of 1:20 on day 56 correlated with the elimination of SARS-CoV-2. Concerning the temporal pattern of appearance and titres of neutralising antibodies in pVNT, variant S:Delta141-4 E484K S494P, which was dominant from day 48 until virus elimination, possessed the strongest immune-escape properties. This highlights the importance of neutralising antibodies against the NTD [41, 43].

While pVNT and sVNT cannot discriminate between antibody classes, this goal was achieved by the in-house EIA. Interestingly, the patientߣs RBD-specific polyclonal immune response between days 49 and 90 appeared to consist solely of IgM. This contrasts with the situation in the immunocompetent host [46-48] and may reflect the gradual recovery of the humoral immune response from the suppressive effect of rituximab administered before SARS-CoV-2 infection. Nevertheless, the emergence of escape mutants and virus elimination provide strong evidence of the neutralising capacity of this immune response. Low avidity and impaired affinity maturation of anti-S antibodies have been suggested to facilitate immune escape in prolonged infection [49]. Due to the lower overall avidity of IgM, the patient’s isolated polyclonal IgM response and the lack of a detectable class switch to IgA or IgG may thus have favoured the emergence of escape mutations, as observed by Truong and colleagues (2021) in a B-ALL patient [50]. On the other hand, engineering high-affinity RBD-specific IgG1 antibodies to IgM has been shown to massively increase the neutralising potency and decrease the resistance of variants [51].

Thus, in a humoral immune response dominated by IgM, intra- and inter-spike binding may become paramount for efficient neutralisation and retaining activity against escape variants as described for bivalent IgG against SARS-CoV-2 and other viruses [52-54]. Concerning the NTD supersite, Fab fragments derived from potently neutralising antibodies have been reported to lose their neutralising activity [55]. This might partially explain the strong immune escape effect of deletion S:Delta141-4 seen in pVNT which does not involve epitopes on the RBD interfering with binding to the hACE2 receptor [56]. In this context, it is noteworthy that during the first episode, only a marginal increase in antibodies against S:S494P or S:E484K S494P, which remained below 30% inhibitory activity, was observed in the sVNT, albeit clearly positive results were obtained in the RBD IgM EIA using identical antigens. Thus, the intrinsic low-avidity binding of IgM to monomeric antigens as compared to the trimeric antigenic structures present in the native S protein may have led to a specific failure in the detection of IgM antibodies in the variant-specific NTD-EIA and in-house sVNT, respectively. In addition, sVNTs may generally exhibit a lower correlation with a full virus neutralisation assay in convalescents as compared to vaccinees [21].

Concerning the period between the onset of antibody-mediated selective pressure and the selection of escape variants during the first episode of infection, it was remarkable that it took almost two weeks for the S:S494P bamlanivimab-resistant variant to emerge. In contrast, the first detection of endogenous, neutralising antibodies against the S:S494P variant coincided with the emergence of escape mutants carrying deletions and substitutions in the NTD and the RBD, respectively. This most likely highlights a considerably improved ability of the S:S494P variant to respond to selective pressure owing to genomic diversity acquired during prolonged infection.

Compared to the first infection, the immune response to the Omicron re-infection differed in several relevant aspects. First, SARS-CoV-2 antibodies elicited by the first infection and/or vaccination were present before re-infection and treatment with sotrovimab. These antibodies reacted in the pVNT with wt S, S:S494P, and S:E484K S494P, however, failed to neutralise S:Delta141-4 E484K S494P and S Omicron BA.1, respectively. In the sVNT, inhibitory antibodies were directed against wt RBD and RBD S494P, whereas RBD E484K S494P or RBD Omicron BA.1-specific inhibitory antibodies were not detected. This reflects the well-known strong immune escape mediated by numerous substitutions in S of the Omicron sublineages [39, 57, 58]. In addition, affinity maturation of the SARS-CoV-2-specific IgG response following the first infection and vaccination may have been impaired [59], resulting in an IgG response with reduced breadth and potency not covering substitutions E484K and Delta141-4.

Second, in contrast to the first episode, re-infection with Omicron BA.1.18 led to a prominent IgA response as seen in the RBD and NTD EIAs. Introducing the substitution S:E340K into the RBD, which interferes with the binding of sotrovimab [22], revealed that Omicron re-infection also elicited a strong reactivity against epitopes on wt RBD and RBD Omicron BA.1 in the sVNT and a strong IgG reaction in the RBD EIA. Since the patient had undergone a B-cell depleting therapy before the first episode but not before re-infection, the serologic response to re-infection with Omicron probably more closely reflected the situation in immunocompetent individuals and may explain the mild, self-limiting course of disease. Interestingly, seroreactivity with the NTD Delta141-4 in the EIA elicited by re-infection with Omicron was lower as compared to wt NTD and NTD Omicron BA.1, respectively. This may hint at additional variant-specific differences in NTD-seroreactivity since S:Delta141-4 observed during the first episode and S:Delta144-5 present in Omicron BA.1 are similar.

There are several limitations to this study. The data presented here only relate to a single patient and can therefore only be applied to the entire group of immunocompromised patients to a limited extent. It can be assumed that the specific previous diseases and forms of therapy used in this case influenced the course of the observed immune response against SARS-CoV-2 variants.

To conclude, our data underline the importance of a quantitatively and qualitatively insufficient endogenous humoral immune response as a trigger of viral intra-host evolution. Specifically, the data presented here emphasise that the onset of an endogenous IgM response in an immunocompromised patient following a prolonged period of virus replication can temporarily create a situation favouring the rapid emergence of escape mutants. The pattern of escape mutations resembles those observed in severely immunosuppressed individuals treated with CCP. Current serological routine assays largely fail to detect and characterise these ongoing, patient-specific serologic responses. Thus, to better evaluate the patient-and variant-specific immune response, it will be crucial to complement monitoring strategies based on genome sequencing in high-risk groups with advanced serological approaches. To this end, our serologic in-house assays have proven as highly sensitive, specific and reproducible tools for the in-depth characterisation of the antibody responses in the immunocompromised host with a prolonged disease course.

## Data Availability

Data are available on reasonable request.

## Acknowledgements

The Institute of Virology is part of the Virus Alliance North Rhine-Westphalia, which is supported by the Ministry of Culture and Science, NRW, Germany. The funders played no role in study design, data collection, analysis and interpretation, the preparation of the manuscript, or the decision to publish.

The plasmid pCG1-SARS-2-S was kindly provided by Prof. Dr. Stefan Pöhlmann (Infection Biology Unit, German Primate Centre, Göttingen, Germany). We thank Gert Zimmer (Institute of Virology and Immunology, Mittelhäusern, Switzerland) for the VSV pseudo-typed virus. We acknowledge support from the Open Access Publication Fund of the University of Muenster.

## Contributors

Conceptualisation: JEK, PRT; data curation: TG, MTH, JEK, AM, MSc, PRT, RV; formal analysis: TG, JEK, AM, MMM, RN, MSc; funding acquisition: SL, AM; investigation: TG, MMM, RN, MLR, MSc; methodology: TG, ERH, JEK, EUL, AM, MLR, MSc; resources (patient samples): MTH, JEK, GL, MSt, PRT, RV; supervision: ERH, EUL, JEK, SL; validation: TG, JEK, SL, EUL, MMM, RN, MLR, MSc; visualisation: JEK, MSc; writing – original draft: TG, JEK, EUL, MSc; writing – review & editing: all authors.

All authors read and approved the final version of the manuscript and agreed to be accountable for all aspects of the work in ensuring that questions related to the accuracy or integrity of any part of the work are appropriately investigated and resolved.

## Data sharing statement

Data are available on reasonable request.

## Declaration of interests

The authors declare no conflict of interest related to this work.

Parts of this study regarding the methodological establishment of the in-house sVNT and EIA were submitted in 2023 by TG to the Medical Faculty of the University of Muenster, Muenster, Germany, as their doctoral thesis entitled “Etablierung serologischer Nachweisverfahren zur Varianten-spezifischen Detektion und Differenzierung von Antikörpern gegen SARS-CoV-2”.

## Ethics Statement

The patient presented in our study has given informed consent to data collection and publication. The ethics committee of the Aerztekammer Westfalen-Lippe, Muenster, Germany, and the University of Muenster has ethically approved the study *Characterisation of the Humoral Immune Response to SARS-CoV-2 in Vaccinees* under the file number 2021-039-f-S. All study participants provided written informed consent.

## Supplementary Material

**Supplementary Table 1a:** List of expression plasmids.

| Vector | Insert |
| --- | --- |
| pCG1-SARS-2-S-Delta1253 | wt S protein with C-terminal 21aa deletion |
| pCG1-SARS-2-S-494P-Delta1253 | wt S protein with C-terminal 21aa deletion and substitution S494P |
| pCG1-SARS-2-S-484K-494P-Delta1253 | wt S protein with C-terminal 21aa deletion and substitutions E484K and S494P |
| pCG1-SARS-2-S-Delta141-4-484K-494P-Delta1253 | wt S protein with C-terminal 21aa deletion, deletion 141-4, and substitutions E484K and S494P |
| pcDNA3.1 SARS-2 Omicron | Omicron BA.1 S protein with C-terminal 21aa deletion |
| pEN-secNL-RBD | secreted RBD fragment of S protein, aa 319 to aa 541, wt sequence, N-terminal NLuc tag |
| pEN-secNL-RBD-E340K | secreted RBD fragment of S protein, aa 319 to aa 541, wt sequence with substitution E340K, N-terminal NLuc tag |
| pEN-secNL-RBD-494P | secreted RBD fragment of S protein, aa 319 to aa 541, wt sequence with substitution S494P, N-terminal NLuc tag |
| pEN-secNL-RBD-484K-494P | secreted RBD fragment of S protein, aa 319 to aa 541, wt sequence with substitutions E484K and S494P, N-terminal NLuc tag |
| pEN-secNL-RBD-Omicron | secreted RBD fragment of S protein, aa 319 to aa 541, Omicron BA.1 sequence, N-terminal NLuc tag |
| pEN-secNL-RBD-Omicron-E340K | secreted RBD fragment of S protein, aa 319 to aa 541, Omicron BA.1 sequence with additional substitution E340K, N-terminal NLuc tag |
| pEN-secNL-NTD | secreted NTD fragment of S protein, aa 15 to aa 307 |
| pEN-secNL-NTD-Delta141-4 | secreted NTD fragment of S protein, aa 15 to aa 307 with deletion 141-4, N-terminal NLuc tag |
| pEN-secNL-NTD-Omicron | secreted NTD fragment of S Omicron BA.1 protein, aa 15 to aa 307 |

**Supplementary Table 1b:**
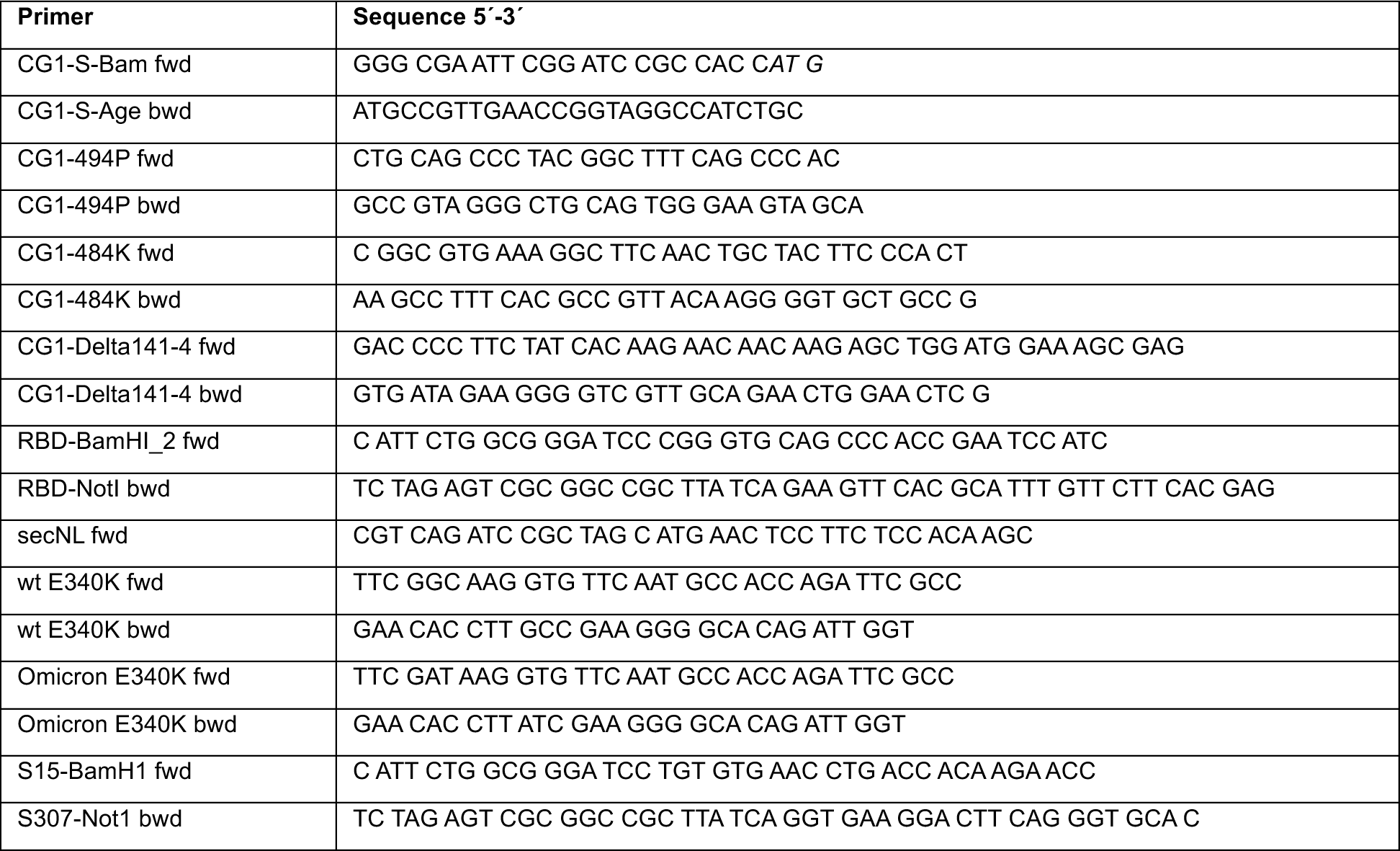
List of primers.

**Supplementary Table 2:**
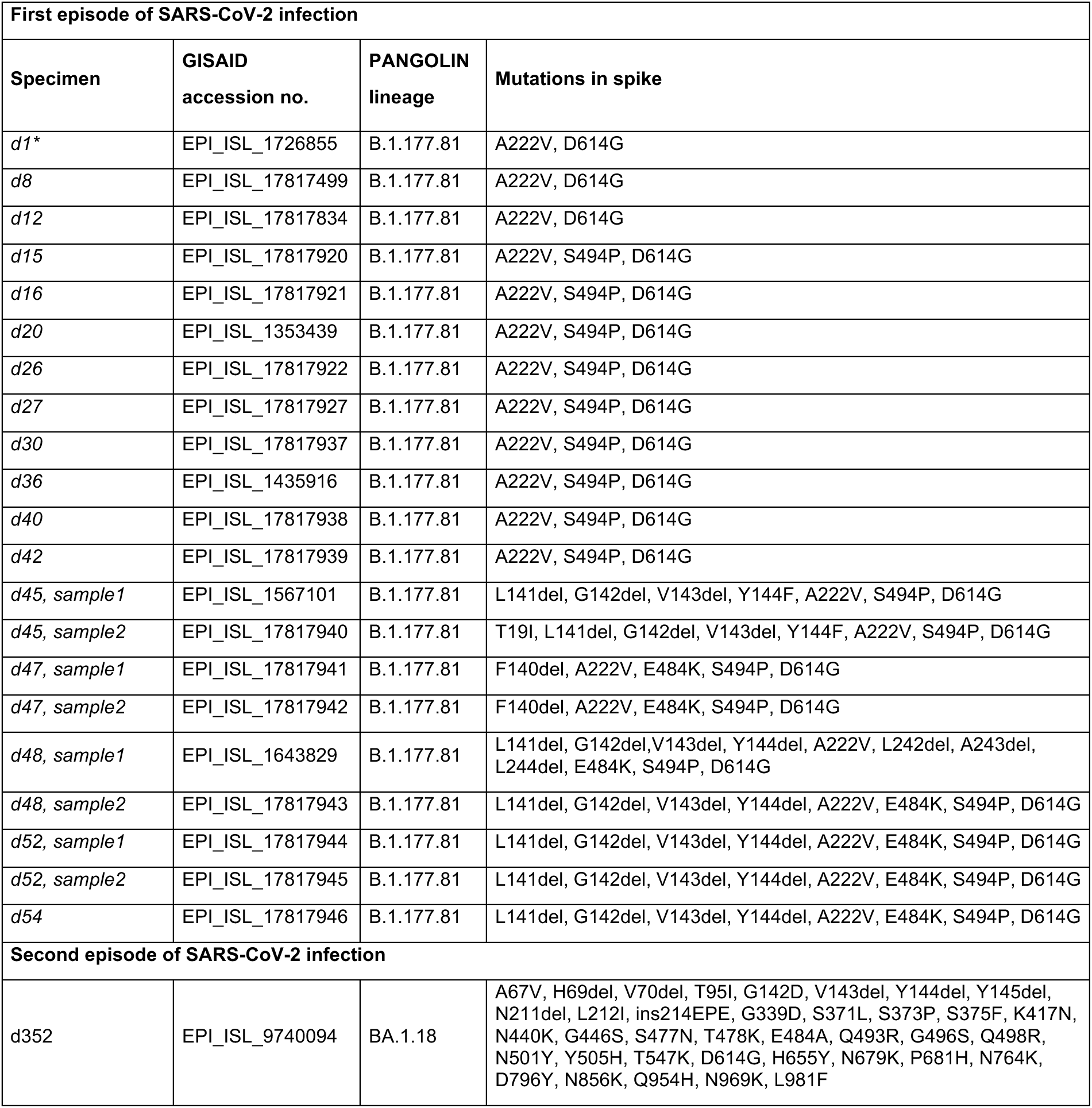
List of specimens, GISAID accession numbers, PANGOLIN lineages and mutations in the spike sequence of virus isolated from the patient.

| First episode of SARS-CoV-2 infection |  |  |  |
| --- | --- | --- | --- |
| Specimen | GISAID accession no. | PANGOLIN lineage | Mutations in spike |
| d1* | EPI_ISL_1726855 | B.1.177.81 | A222V, D614G |
| d8 | EPI_ISL_17817499 | B.1.177.81 | A222V, D614G |
| d12 | EPI_ISL_17817834 | B.1.177.81 | A222V, D614G |
| d15 | EPI_ISL_17817920 | B.1.177.81 | A222V, S494P, D614G |
| d16 | EPI_ISL_17817921 | B.1.177.81 | A222V, S494P, D614G |
| d20 | EPI_ISL_1353439 | B.1.177.81 | A222V, S494P, D614G |
| d26 | EPI_ISL_17817922 | B.1.177.81 | A222V, S494P, D614G |
| d27 | EPI_ISL_17817927 | B.1.177.81 | A222V, S494P, D614G |
| d30 | EPI_ISL_17817937 | B.1.177.81 | A222V, S494P, D614G |
| d36 | EPI_ISL_1435916 | B.1.177.81 | A222V, S494P, D614G |
| d40 | EPI_ISL_17817938 | B.1.177.81 | A222V, S494P, D614G |
| d42 | EPI_ISL_17817939 | B.1.177.81 | A222V, S494P, D614G |
| d45, sample1 | EPI_ISL_1567101 | B.1.177.81 | L141del, G142del, V143del, Y144F, A222V, S494P, D614G |
| d45, sample2 | EPI_ISL_17817940 | B.1.177.81 | T19I, L141del, G142del, V143del, Y144F, A222V, S494P, D614G |
| d47, sample1 | EPI_ISL_17817941 | B.1.177.81 | F140del, A222V, E484K, S494P, D614G |
| d47, sample2 | EPI_ISL_17817942 | B.1.177.81 | F140del, A222V, E484K, S494P, D614G |
| d48, sample1 | EPI_ISL_1643829 | B.1.177.81 | L141del, G142del, V143del, Y144del, A222V, L242del, A243del, L244del, E484K, S494P, D614G |
| d48, sample2 | EPI_ISL_17817943 | B.1.177.81 | L141del, G142del, V143del, Y144del, A222V, E484K, S494P, D614G |
| d52, sample1 | EPI_ISL_17817944 | B.1.177.81 | L141del, G142del, V143del, Y144del, A222V, E484K, S494P, D614G |
| d52, sample2 | EPI_ISL_17817945 | B.1.177.81 | L141del, G142del, V143del, Y144del, A222V, E484K, S494P, D614G |
| d54 | EPI_ISL_17817946 | B.1.177.81 | L141del, G142del, V143del, Y144del, A222V, E484K, S494P, D614G |
| Second episode of SARS-CoV-2 infection |  |  |  |
| d352 | EPI_ISL_9740094 | BA.1.18 | A67V, H69del, V70del, T95I, G142D, V143del, Y144del, Y145del, N211del, L212I, ins214EPE, G339D, S371L, S373P, S375F, K417N, N440K, G446S, S477N, T478K, E484A, Q493R, G496S, Q498R, N501Y, Y505H, T547K, D614G, H655Y, N679K, P681H, N764K, D796Y, N856K, Q954H, N969K, L981F |

## Notes

### Competing Interest Statement

The authors have declared no competing interest.

### Author Declarations

The patient presented in our study has given informed consent to data collection and publication. The ethics committee of the Aerztekammer Westfalen-Lippe, Muenster, Germany, and the University of Muenster has ethically approved the study Characterisation of the Humoral Immune Response to SARS-CoV-2 in Vaccinees under the file number 2021-039-f-S. All study participants provided written informed consent.

